# Maternal adverse childhood experiences on child growth and development in rural Pakistan: an observational cohort study

**DOI:** 10.1101/2023.02.10.23285752

**Authors:** Esther O. Chung, Elissa Scherer, Katherine LeMasters, Lisa Bates, Ashley Hagaman, Brooke S. Staley, Lauren Zalla, Siham Sikander, Joanna Maselko

## Abstract

Maternal adverse childhood experiences (ACEs) have been associated with negative impacts on children. However, not all types or levels of adversity are similarly deleterious and research from diverse contexts is needed to better understand why and how intergenerational transmission of adversity occurs. We examined the role of maternal ACEs on child growth and development at 36 months postpartum in rural Pakistan. We used data from 877 mother-child dyads in the Bachpan Cohort, a birth cohort study. Maternal ACEs were captured using an adapted version of the ACE-International Questionnaire. Outcomes included child growth, fine motor and receptive language development, and socioemotional and behavioral development at 36 months of age. To estimate the associations between maternal ACEs and child outcomes, we used multivariable generalized linear models with inverse probability weights to account for sampling and loss to follow-up. Over half of mothers in our sample (58%) experienced at least one ACE. Emotional abuse, physical abuse, and emotional neglect were the most commonly reported. We found null relationships between the number of maternal ACEs and child growth. Maternal ACEs were associated with higher fine motor and receptive language development and worse socioemotional and behavioral outcomes. Maternal ACE domains had similarly varying relationships with child outcomes. Our findings highlight the complexity of intergenerational associations between maternal ACEs and children’s growth and development. Further work is necessary to examine these relationships across cultural contexts and identify moderating factors to mitigate potential negative intergenerational effects.

## Introduction

Adverse childhood experiences (ACEs) refer to severely stressful exposures or experiences that occur in childhood, such as abuse, neglect, violence between caregivers, and peer or community violence [1]. The effects of ACEs start early in life and continue throughout the lifecourse, beginning with delayed child development and progressing to poor psychological and physical health outcomes in adulthood [2, 3].

ACEs have been linked to numerous adult health outcomes such as cardiometabolic disease, cancer, mortality [4–6], and several negative psychological outcomes including depression, post-traumatic stress disorder, anxiety, and suicide [7–10]. Severe early life stressors can lead to observable changes in brain structure and function, which has potentially permanent effects on development [11, 12], and can alter physiological systems, such as the stress response axes, immune functioning and inflammation [13].

While there is considerable evidence of the harmful impact of accumulated ACEs across an individual’s life course, intergenerational effects are an understudied area of concern. The intergenerational transmission of maternal childhood adversity may occur through biological embedding during pregnancy as well as through maternal mental health and parenting related pathways [14]. For example, stress-induced epigenetic alterations during a mother’s childhood can alter the maternal-placental-fetal endocrine, immune, and inflammatory stress biology. This, in turn, may affect the fetal brain and ultimately, the child’s physical, cognitive, and socioemotional developmental trajectories. Maternal ACEs may also impact the quality of the postnatal environment [14] and, indeed, maternal anxiety, depression, and parenting practices are key mediators between maternal ACEs and child socioemotional development [15–18].

In relation to parental ACEs and early childhood development, the most studied domain is socioemotional development. Worse socioemotional functioning, evident through greater externalizing and internalizing problems, has been shown to exist for children born to mothers with greater exposure to ACEs in high-income countries [15,19–21]. These socioemotional problems include higher anxiety, aggression, hyperactivity, and negative affect in the first three years of life [9,22,23].

Less research has been conducted examining the impact of maternal childhood adversity and the next generation’s physical growth and cognitive development. Of the existing research, maternal ACEs have been linked to low birth weight and shorter gestational age [24], lower overall development at 12 months [25], lower parental-rated physical health at 18 months [26], and decreased problem solving, gross motor, fine motor, and communication skills at 24 months [27]. All of these studies were conducted in the United States (US) or Canada, and few studies have examined the intergenerational effects of maternal ACE exposure in low- and middle- income countries (LMICs). Addressing ACEs in LMICs is critically important for several reasons. Adversity in childhood is more prevalent in LMICs, resources to address ACEs in LMICs are limited, and the majority of the world’s population resides in LMICs [28]. Finally, understanding the intergenerational effects of maternal ACEs across cultural contexts will help to identify vulnerable populations for intervention. To our knowledge, we are only aware of three studies assessing the intergenerational effects of maternal ACEs in LMICs. In one recent study, researchers reported that maternal ACEs were associated with poor fetal attachment during pregnancy across eight LMICs; however, the effects varied across cultural contexts [29]. Researchers reported positive effect estimates of maternal ACEs on fetal attachment in Jamaica, Pakistan, Philippines, and South Africa, no effect in Sri Lanka, and negative effect estimates in Ghana, Romania, and Vietnam. Two other studies found maternal ACEs were associated with worse socioemotional development among older children (aged 2-18 years) in Kenya [17, 18]. The heterogeneity of results from these studies underscore the importance of studying the intergenerational effects of maternal childhood adversity across diverse contexts. These studies also did not investigate how maternal ACEs affected early child growth and development. Therefore, the present study examined the role of maternal ACEs in child growth and development at 36 months postpartum in rural Pakistan.

## Methods

### Study Population and Participants

We used data from the Bachpan Cohort, a birth cohort study based in Kallar Syedan, a rural subdistrict in Rawalpindi, Punjab Province [30, 31]. Kallar Syedan has a population of roughly 200,000 people and the average household size is six individuals [32]. Literacy rates for men and women in rural Punjab province are 70% and 53%, respectively. Women in our sample had higher educational attainment compared to the Demographic and Health Survey (DHS) rates in rural areas of the Punjab province (50% in our sample achieved secondary school or higher vs. 20% in the DHS) [33].

Pregnant women in their third trimester were invited for depression screening from 2014-2016. Women who screened positive for depression were invited to participate in the trial portion of the study, and a random sample of a third of women who screened negative for depression were asked to participate in the cohort only portion, which created a population-representative sample. Mother-child dyads were enrolled in their third trimester and followed up at 3, 6, 12, 24, and 36 months postpartum. Of the 1,154 women assessed at baseline, 265 were lost to follow-up by 36 months and 12 were missing outcome measurements, resulting in 877 mother-child dyads with complete data.

### Exposure

We measured maternal ACEs using an adapted version of the ACE-International Questionnaire (ACE-IQ) [34], a self-reported retrospective measure that has been validated in other low-resource contexts (S1 Table) [35, 36]. The ACE-IQ was adapted from the original ACE measure developed in the US [37] to include items on peer violence, exposure to collective violence and witnessing community violence. Due to potential risks to the participant and expected underreporting, we removed the sexual abuse questions. At 36 months postpartum, mothers retrospectively reported their experiences of 12 adverse exposures in childhood.

We operationalized maternal ACEs in multiple ways. We created a summed score ranging from 0-12, a categorical variable with the number of experiences (0, 1, 2, 3, 4 or more), and domain-specific indicators for neglect (emotional and physical), family psychological distress (alcohol and/or drug abuser in the household; incarcerated household member; someone depressed, mentally ill, institutionalized or suicidal), home violence (physical abuse; emotional abuse; household member treated violently), and community violence (bullying; community violence; collective violence). For each domain-specific indicator, women received a “Yes” if they experienced any of the ACEs within each domain. Twenty women responded “Do not remember” to some ACE questions and these were recoded as “No”.

### Outcomes

Our main outcomes were child growth and development (fine motor, receptive language, and socioemotional) at 36 months. We used the World Health Organization’s standards to measure physical growth z-scores using length-for-age (LAZ), weight-for- age (WAZ), and weight-for-length (WLZ) [38]. We measured fine motor and receptive language development using the Bayley Scales of Infant and Toddler Development [39]. Scaled scores were calculated using a reference population in the US by the child’s age group; scores range from 1-19 with a mean of 10 and standard deviation of three.

Socioemotional development was captured using the Ages and Stages Questionnaire: Socioemotional (ASQ:SE) [40]. The ASQ:SE is a 30-item parent-reported measure, ranging from 0-270, with higher scores indicating worse child socioemotional development. Behavioral outcomes were assessed using the parent-reported Strengths and Difficulties Questionnaire (SDQ) [41]. The SDQ covers 25 questions in five domains: hyperactivity, emotional, conduct, and peer problems, and pro-social behaviors. A total score is calculated by summing the first four domains and ranges from 0-40, with higher scores indicating worse behavioral outcomes.

### Confounders

We included the following baseline confounders informed by a directed acyclic graph (DAG) [42, 43] to estimate the total effect of maternal ACEs on early child growth and development: maternal age, education, and family history of mental illness. We aimed to estimate the total effect of maternal ACEs on child outcomes; therefore, we did not control for potential variables, such as maternal depression or child health, which we identified as mediators using the DAG. We used maternal education as a proxy for maternal childhood socioeconomic status and operationalized it as a categorical variable (none, primary or middle school, secondary education or higher). We asked mothers to report if anyone in her immediate natal family had an existing mental illness and used it as a proxy for the mother’s family history of mental illness.

### Statistical Analysis

We constructed multivariable generalized linear models to assess the relationship between maternal ACEs and child growth and development. In addition to confounders, we also included child gender, trial arm, and assessor in all models to improve precision [44]. We used cluster robust standard errors to account for clustering by village, and cluster-specific sampling weights to account for unequal sampling probabilities by baseline depression. In a given cluster, all non-depressed women were weighted by the inverse of their sampling fraction (i.e., the proportion that were screened for depression at recruitment and subsequently enrolled), and depressed women were each given a weight of 1. Therefore, non-depressed women were upweighted to account for their lower probability of selection into the study.

To account for informative censoring between baseline and 36 months postpartum, we used stabilized inverse probability of censoring weights (IPCW) [45]. IPCW are estimated as the inverse of the probability that a woman was not censored at 36 months, based on observed characteristics. Women who were not loss to follow-up were upweighted to represent those who were lost to follow-up. In the IPCW model, we included baseline confounders (maternal age, education, and family history of mental illness) and baseline characteristics associated with censoring using a p-value <0.10 (maternal depression, child’s grandmother co-residence, number of people per room, and nuclear vs. joint family), with household asset scores included to increase precision [46]. IPCW were stabilized by using the marginal probability of being observed in the numerator. Sampling weights and IPCW were multiplied to create a final weight used in all models [47]. The final weight was also used to estimate means and frequencies in descriptive tables; numbers were unweighted. Analyses were conducted in Stata 14 and R.

### Ethics approval

The Bachpan Cohort study received ethical approval from the institutional review boards at the Human Development Research Foundation (IRB/1017/2021), Duke University, and the University of North Carolina at Chapel Hill (#20-1433). Written informed consent, or witnessed informed consent if the participant was illit- erate, was obtained before study participation.

## Results

### Descriptive statistics

There were 877 women and children in our analytic sample (Table 1). After applying weights, on average, women were 27 years old and half had completed secondary school education or higher at baseline. Roughly 28% of mothers were diagnosed with depression at baseline and 9% reported having a family history of mental illness. The majority of households were joint or extended families and 68% of children were co-residing with a grandmother. Almost half of the children were female and mean LAZ and WAZ were -0.99 (SD=1.10) and -0.96 (SD=0.99), respectively. Mean fine motor and receptive language scaled scores were slightly higher than the average of 10 (10.43 [2.79]; 11.45 [4.03], respectively). Mean ASQ-SE and SDQ scores were 38.83 (SD=17.03) and 14.02 (SD=6.30), respectively.

**Table 1.**
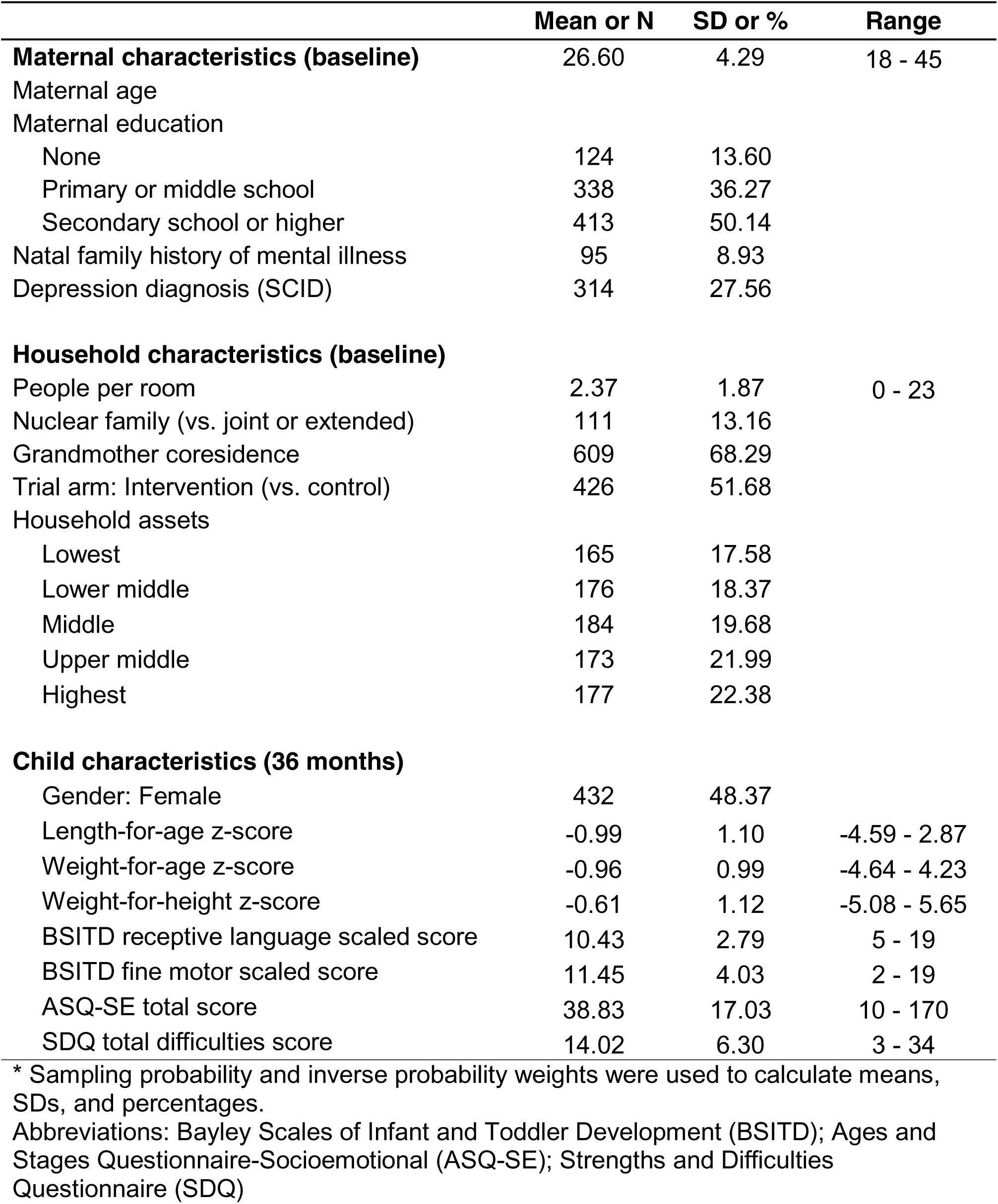
Sample characteristics, Bachpan Cohort, Pakistan, n=877.

After applying weights, over 58% of women (n=512) experienced at least one ACE (Table 2). Among the 12 ACE questions, emotional abuse (n=290, 32%), physical abuse (n=206, 23%), emotional neglect (n=131, 15%), and witnessing a household member being treated violently (n=128, 15%) were the most common. With respect to ACE domains, more than one in three women were exposed to violence in their childhood home (n=344, 39%) and one in five experienced emotional or physical neglect during their childhood (n=170, 19%). Roughly 16% (n=141) experienced family psychological distress. Community violence was far less common (n=63, 6.9%).

**Table 2.** Maternal Adverse Childhood Experiences (n=877)*

|  | **N** | **%** |
| --- | --- | --- |
| **Number of ACEs** |  |  |
| None | 363 | 41.49 |
| 1 | 235 | 26.86 |
| 2 | 137 | 15.66 |
| 3 | 81 | 9.26 |
| 4 or more | 59 | 6.74 |
| **Neglect** |  |  |
| Physical neglect | 49 | 5.60 |
| Emotional neglect | 131 | 14.97 |
| **Family psychological distress** |  |  |
| One or no parents, parental separation or divorce | 96 | 10.97 |
| Alcohol/drug abuser in the household | 23 | 2.63 |
| Someone chronically depressed, mentally ill | 21 | 2.40 |
| Incarcerated household member | 16 | 1.83 |
| **Home violence** |  |  |
| Household member treated violently | 128 | 14.63 |
| Physical abuse | 206 | 23.54 |
| Emotional abuse | 290 | 33.14 |
| **Community violence** |  |  |
| Bullying | 12 | 1.37 |
| Community violence | 59 | 6.74 |
| Collective violence | 4 | 0.46 |
| **ACE domains\*\*** |  |  |
| Neglect | 170 | 19.43 |
| Family psychological distress | 141 | 16.11 |
| Home violence | 344 | 39.31 |
| Community violence | 63 | 7.20 |
|  | **Mean** | **SD** |
| **Total number of ACEs** (range: 0-10) | 2.37 | 1.38 |

\* Sampling probability weights and inverse probability weights were used.
\*\* Domains were binary categories describing experience of any individual ACEs.

We highlight the effects of maternal ACEs on child growth and development below. Full estimates and precision are presented in S2 and S3 Tables.

### Growth outcomes

Maternal ACEs and child growth were not strongly associated (Fig 1, S2 Table); our estimates were noisy, however, we found some patterns. There were small, negative relationships between maternal ACEs and child LAZ (Fig 1A). We found a negative trend with child LAZ as maternal ACEs increased with four or more maternal ACEs were associated with a 0.13 SD decrease in child LAZ (95% CI: -0.42, 0.15).

**Fig 1.**
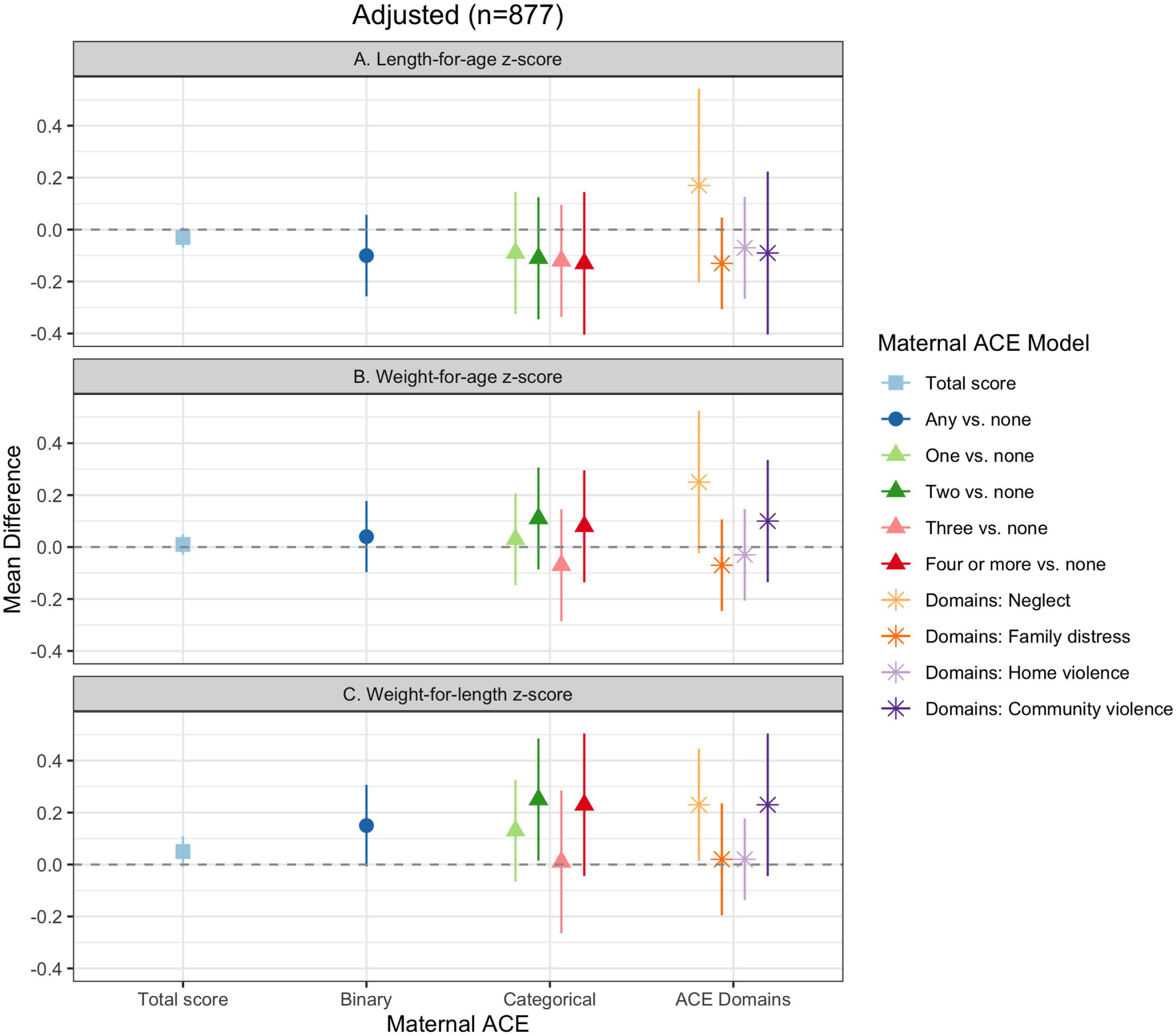
Maternal ACEs and child growth, Bachpan Cohort, Pakistan (n=877). We used weighted generalized linear models with cluster robust standard errors. Sampling and inverse probability censoring weights were combined. All models controlled for baseline maternal age, maternal education, trial arm, assessor, and child gender.

Except for the neglect domain, family distress, home violence, and community violence had negative associations with child LAZ (Fig 1A). We found largely null relationships between maternal ACEs and child WAZ (Fig 1B); however, neglect was positively associated with WAZ (0.25, 95% CI: -0.04, 0.53). Maternal ACEs and WLZ were slightly positively associated (Fig 1C). Any maternal ACE compared to none was associated with a 0.15 SD increase in WLZ (95% CI: -0.02, 0.32). There was no clear stepwise trend between incremental ACEs and WLZ; however, four or more maternal ACEs were associated with a 0.23 SD higher WLZ (95% CI: -0.06, 0.52). Maternal childhood experience of neglect and community violence were also associated with a 0.23 increase in child WLZ (95% CI: 0.01, 0.45; -0.05, 0.52, respectively).

### Fine motor and receptive language outcomes

We found positive associations between maternal ACEs and children’s fine motor and receptive language scores (Fig 2, S3 Table). Any maternal experience of ACEs was associated with both higher fine motor and receptive language scores compared to no maternal ACE. There was not a clear stepwise trend in incremental maternal ACE exposure and fine motor scores (Fig 2A). In contrast, we saw a small stepwise trend where, compared to no maternal ACEs, experience of two, three, and four or more ACEs was associated with incremental increases in children’s receptive language scores (Fig 2B). Maternal experiences of neglect and family distress during her childhood were associated with higher child fine motor and receptive language scores (Fig 2). Home violence was not associated with fine motor or receptive language scores. Community violence was associated with decreased fine motor scores and increased receptive language scores; however, these estimates were imprecise with large confidence limit differences (Fig 2).

**Fig 2.**
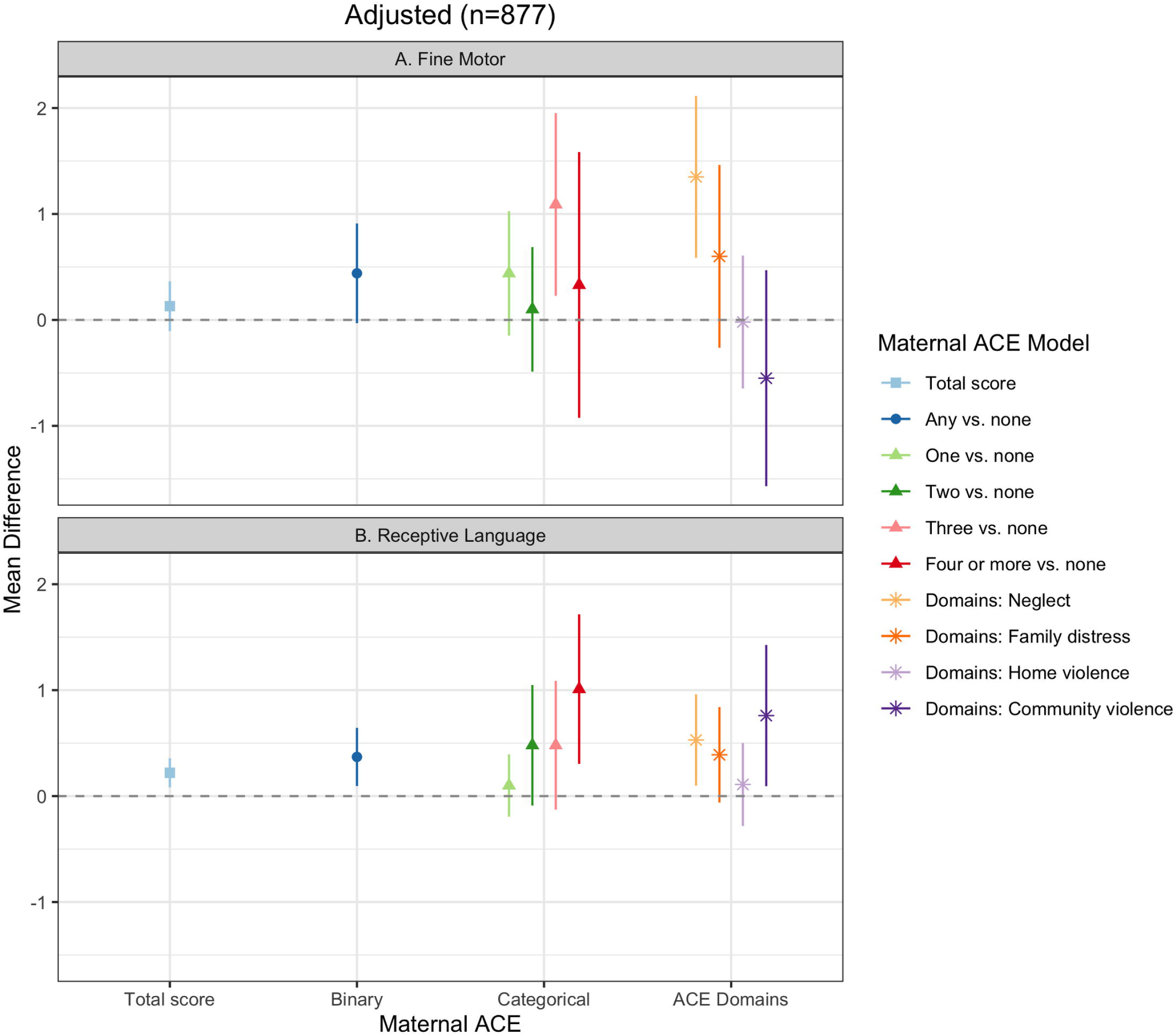
Maternal ACEs and child fine motor and receptive language, Bachpan Cohort, Pakistan (n=877). We used weighted generalized linear models with cluster robust standard errors. Sampling and inverse probability censoring weights were combined. All models controlled for baseline maternal age, maternal education, trial arm, assessor, and child gender.

### Socioemotional outcomes

Maternal ACEs were associated with worse child socioemotional and behavioral development; higher ASQ:SE and SDQ scores suggest greater socioemotional problems (Fig 3, S3 Table). Compared to children of mothers who experienced no ACE, children whose mothers experienced any ACE scored 3.30 points higher on the ASQ:SE (95% CI: 0.98, 5.62). We found a similar effect on SDQ; however, the mean difference was substantially smaller (Fig 2B, MD=0.59, 95% CI: -0.27, 1.44). There was an increasing trend between incremental maternal ACE exposure on ASQ:SE, but not for SDQ (Fig 3A and 3B). Maternal experience of all four domains (neglect, family distress, home violence, and community violence) were associated with higher ASQ:SE scores; however, our confidence intervals were wide with confidence limit differences ranging from 3.95 to 7.43 (Fig 3A). For SDQ, maternal childhood experience of home violence was associated with higher scores, while maternal childhood experience of neglect and family distress had small negative associations with child SDQ.

**Fig 3.**
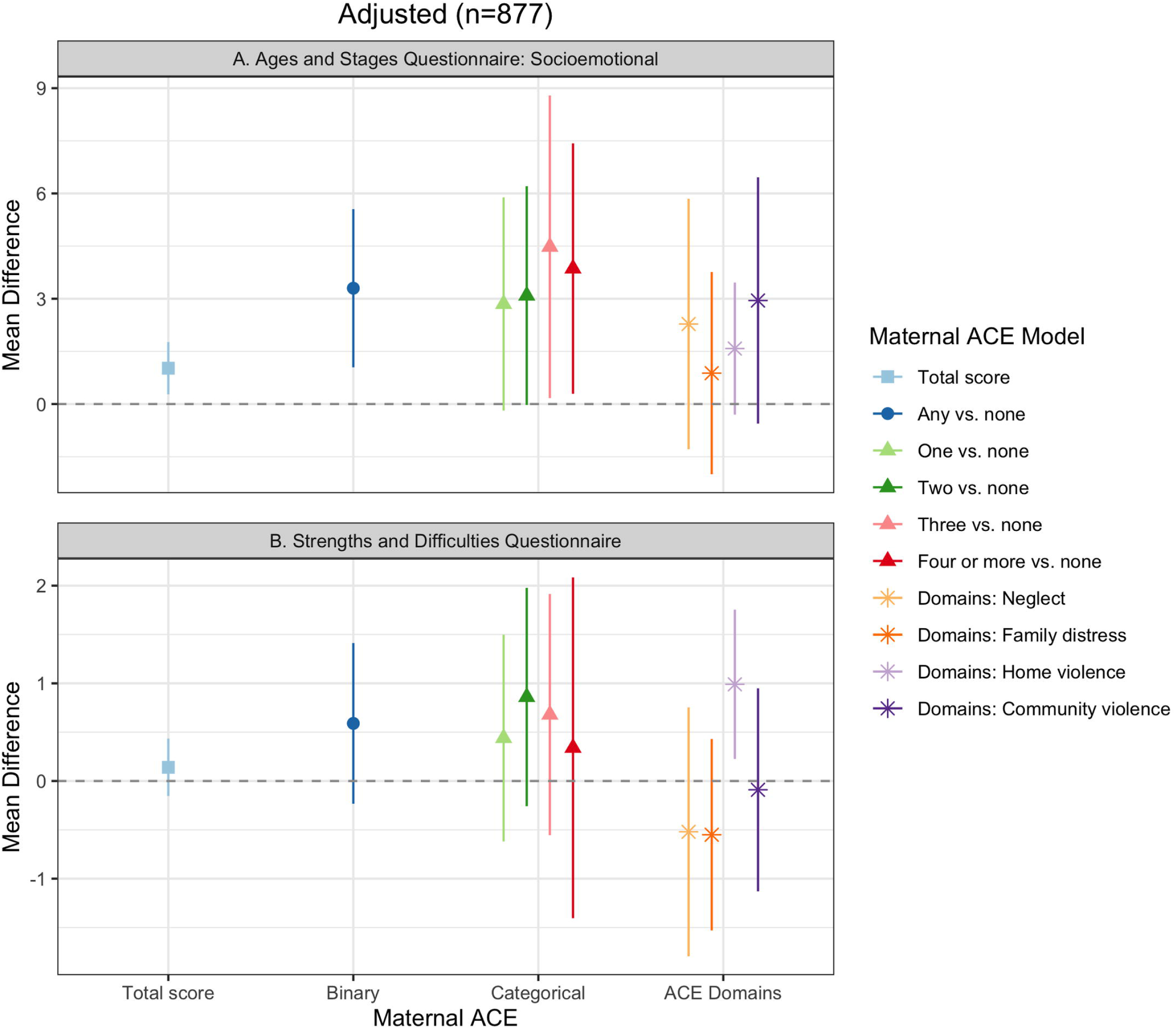
Maternal ACEs and child socioemotional and behavioral outcomes, Bachpan Cohort, Pakistan (n=877). We used weighted generalized linear models with cluster robust standard errors. Sampling and inverse probability censoring weights were combined. All models controlled for baseline maternal age, maternal education, trial arm, assessor, and child gender.

## Discussion

The goal of our analysis was to estimate the effects of maternal ACEs on her child’s development at 36 months of age. Over half of the mothers in our sample in rural Pakistan experienced at least one ACE. Maternal ACEs were not associated with child growth. We found an unexpected relationship between maternal ACEs and child fine motor and receptive language development, where maternal childhood experience of adversity was associated with more positive child development. Maternal ACEs were associated with worse child socioemotional and behavioral outcomes.

Mothers most commonly reported experiences of emotional abuse, physical abuse, emotional neglect, or seeing a household member treated violently during their childhood. The prevalence of any maternal ACEs in our sample (58%) was lower than in other LMICs using the same measure (ACE-IQ, roughly 80% in Saudi Arabia, Vietnam, Tunisia, and Lebanon) [48]. Other studies in Pakistan have also reported lower levels of ACEs. For example, one study of Pakistani university students found the mean number of ACEs among women was 0.40 (SD=0.92) [49], compared with our mean of 2.37 (SD=1.38). The authors of that study emphasized that in a collectivist and conservative context like Pakistan, the role of the family is critical even throughout adulthood, making it especially difficult to disclose sensitive topics, such as child maltreatment and family distress. Another study of pregnant women in Pakistan also reported a similar mean number of maternal ACEs in Pakistan (3.07, SD=2.37) as our study (2.37, SD=1.38) [29]. This study also found a wide range of mean ACEs across eight LMICs (from 2.54 in Vietnam to 6.42 in South Africa) [29], thus underscoring the importance of examining diverse cultural and contextual factors when assessing ACEs. Specific to measurement, another reason for the lower prevalence of ACEs in our study may be due to our exclusion of the sexual abuse questions, which was done given potential risks to participants and expected underreporting of disclosure. Overall, cross-country variation in mean number of reported ACEs may therefore be due to both differential reporting and true variation in ACE exposures.

In our study, maternal ACEs were not associated with child growth outcomes at 36 months. If a key mechanism through which maternal ACEs influence growth is through the intrauterine environment in pregnancy, it is possible that any such effects would be observed closer to birth. The potential negative effects of maternal ACEs on children’s physical development could have also been remedied between birth and 36 months through moderating factors such as improved nutrition. During the first three years of life, other external factors might have stronger influences on child growth. In low-resource contexts, other forms of early life adversity not captured by ACEs, such as extreme poverty, may have a greater effect on children’s physical development than maternal ACEs.

Notably, maternal ACEs were associated with better children’s fine motor and receptive language scores. There is limited literature examining the intergenerational transmission of maternal ACEs on children’s overall developmental outcomes. Existing research from high-income countries are mixed. In two studies from the US and Canada, maternal ACEs were associated with worse overall development and greater risk of developmental delay [25, 27], while another US study found no relationship between maternal ACEs and child development [50]. We found no prior study investigating maternal ACEs and overall child development in LMICs. While more research is needed to replicate these findings, the idea of post-traumatic growth may, in theory, explain a positive relationship between maternal ACEs and better child development. Post-traumatic growth is defined as a positive psychological change following traumatic events [51–53]. Prior work has found that individuals who experience past adversity have increased empathy and altruism for others [54–56]. In support of this hypothesis, Brown et al (2021) found a positive effect of maternal ACEs on fetal attachment among Pakistani mothers, although they did not examine child development. Importantly, the exact mechanisms through which this would be linked to improved developmental outcomes remains unknown. Assessing the ways in which post-traumatic growth among mothers who experienced ACEs affects health and social behaviors across the lifecourse would be invaluable.

In contrast to the findings with overall development, maternal ACEs in our sample were associated with worse child socioemotional development and behavioral outcomes. This finding is more consistent with the existing literature. In a systematic review, authors found maternal ACEs were associated with child externalizing behavior problems and the majority of included studies found relationships with child internalizing problems [57]. Similar results have been found in Kenya among older aged children (2- 18 years) [17, 18]. These links can potentially be explained through both biological and psychological pathways. Potential biological mechanisms suggest maternal experience of childhood adversity may influence maternal HPA-axis functioning during pregnancy, leading to changes in the gestational environment and the development of fetal stress response systems [58, 59]. The hormonal changes during pregnancy may themselves, in turn, influence the woman’s mental health and health behaviors [60, 61], which can then affect infant stress regulation and behavioral difficulties [62]. Psychosocial mechanisms posit that maternal ACEs can lead to poor attachment and development of healthy relationships between mothers and children [57]. Mothers who experience childhood trauma may be predisposed to mental health conditions such as anxiety or depression, and poor maternal mental health has been linked to increased parenting stress, impaired mother-child interactions, and harsher maternal parenting practices [63]. Furthermore, in the context of LMICs, economic and parenting resources may be scarce and this may exacerbate stress in the home environment. This, in turn, may lead to suboptimal caregiving (lack of warmth and responsiveness) and ultimately, worse child socioemotional development and behavior outcomes.

Finally, maternal ACE domains (Neglect, Family distress, Home violence, and Community violence) had varying relationships with child development. Maternal childhood exposure to neglect was associated with better child growth, fine motor, and receptive language development, but worse child socioemotional development.

Maternal childhood experience of family distress, home violence, and community violence had varied relationships with child outcomes. It is important to note that deprivation (absence of cognitive and social inputs) and threat (presence of threatening experiences) likely have distinct influences on neural development [64], and research has shown differential impacts on cognitive and emotional outcomes in children as well as pregnant women [65–67]. As previously mentioned, post-traumatic growth and compensatory parenting practices may help to explain the positive relationships of past maternal neglect on her own child’s outcomes [51,52,68]. Positive experiences, such as supportive relationships and prosocial activities, can help to mitigate the impact of ACEs by serving as a promotive resilience factor [68]. Prior research demonstrates that adults who had positive childhood experiences are more likely to have resilient functioning and more compensatory behaviors (e.g. less harsh parenting attitudes, greater affection) even if they experienced ACEs [69–73]. Mothers who experienced neglect in their childhood and had positive experiences concurrently or later in childhood may develop resiliency and in turn provide more resources and improved parenting for their children. Future research in LMICs should focus on identifying differential impacts of ACE domains on the next generation and examine whether and how positive experiences can mitigate the intergenerational transmission.

Although such explorations are beyond the scope of the current paper, important factors, such as maternal depression and parenting practices, may mediate the relationship between maternal ACEs and child outcomes. Previous work from our group demonstrated that ACEs were associated with greater depressive symptoms and major depressive episodes [74]. Studies in Kenya found that maternal mental health mediated the relationship between maternal ACEs and child mental health problems [17, 18]. A study among pregnant mothers across eight LMICs found prenatal depression fully mediated the effect of maternal ACEs on fetal attachment using the full sample [29].

However, at the country-level, direct effects varied; in Vietnam, maternal ACEs directly and negatively affected fetal attachment and maternal prenatal depression did not mediate the relationship [29]. Other caregivers, such as fathers and grandmothers, are also important to consider. Their contributions may buffer any negative effects of maternal ACEs on children by improving maternal social support.

### Strengths and limitations

Our study had several strengths. We used standard, validated measures of child outcomes that have been previously used in our study context. Moreover, we estimated the total effect of maternal ACEs on the next generation’s development as opposed to controlling for potential mediators, such as adult socioeconomic status or depression status, which may lead to biased estimates. Our study also had limitations that require discussion. First, although we used the ACE-IQ to capture maternal ACEs, it is a retrospective measure, which is vulnerable to recall bias. In the context of this study, it may be that depressed women are more likely to report ACEs in order to understand their depressive symptoms, resulting in potential differential misclassification. Moreover, the ACE-IQ may not fully measure adversities during childhood in this context. Second, while we controlled for maternal childhood SES and maternal family history of mental illness, we used proxies (maternal education and natal family mental illness history, respectively), and residual confounding is possible. Third, 265 women and children were lost to follow-up at 36 months postpartum; however, we used stabilized IPCW to account for informative censoring. When we excluded these weights, the estimates and precision did not substantially change, indicating that our results were not sensitive to weighting (S4 Table). Finally, there may be unmeasured confounders that affect this relationship, such as how mothers were raised (i.e., the caregiving practices of the mother’s parents).

## Conclusion

Our findings suggest the intergenerational transmission of childhood adversity is complex and may differentially influence physical, cognitive, and socioemotional domains of child development. To improve child health and development, it is necessary to understand maternal life histories, including her childhood experiences, rather than only capturing her experiences during pregnancy and postpartum. Future work should examine the intergenerational relationship of childhood adversity across cultural contexts in order to understand specific risk and protective factors and develop tailored interventions. Identifying moderating factors, such as promoting maternal responsive caregiving and improving maternal social support via other caregivers, can help inform strategies to disrupt the intergenerational cycle of adversity that is responsive to traumatic histories.

## Data Availability

Data are available upon reasonable request from the senior author, Joanna Maselko. Data are not publicly available due to ongoing data collection however, data will be released when the Bachpan cohort study is completed.

## Acknowledgements

The authors are deeply grateful to the women, children, and communities of the Bachpan cohort. We would also like to thank the team at the Human Development Research Foundation (HDRF) including Rakshanda Liaqat, Tayyiba Abbasi, Maria Sharif, Samina Bilal, Quratul-Ain, Anum Nisar, Amina Bibi, Shaffaq Zufi- qar, Sonia Khan, Ahmed Zaidi, Ikhlaq Ahmad, and Najia Atif for their meaningful contributions to the study’s design and implementation. We also thank the larger Bachpan and SHARE CHILD study teams. We are grateful for Paul Zivich’s advice and guidance for implementing inverse probability weights.

## Supporting Information

**S1 Table. Adapted Adverse Childhood Experiences International Questionnaire (ACEs-IQ)**

**S2 Table. Maternal ACEs and child growth, Bachpan Cohort, Pakistan (n=877)**

**S3 Table. Maternal ACEs and child development, Bachpan Cohort, Pakistan (n=877)**

**S4 Table. Maternal ACEs and child growth and development without censoring weights, Bachpan Cohort, Pakistan (n=877**

