## Supplemental Files for "Maternal adverse childhood experiences on child growth and development in rural Pakistan: an observational cohort study"

**S1 Table. Adapted Adverse Childhood Experiences International Questionnaire (ACEs-IQ)**

**Neglect**

*Physical neglect*

Did your parents/guardians not give you enough food even when they could easily have done so? OR Were your parents/guardians too drunk or intoxicated by drugs to take care of you? OR Did your parents/guardians not send you to school even when it was available?

*Emotional neglect*

Did your parents/guardians understand your problems and worries? OR Did your parents/guardians really know what you were doing with your free time when you were not at school or work?

**Family psychological distress**

*One or no parents, parental separation or divorce*

Were your parents ever separated or divorced? OR Did your mother, father or guardian die?

*Alcohol/drug abuser in the household*

Did you live with a household member who was a problem drinker or alcoholic, or misused street or prescription drugs?

*Someone chronically depressed, mentally ill*

Did you live with a household member who was depressed, mentally ill or suicidal?

*Incarcerated household member*

Did you live with a household member who was ever sent to jail?

**Home violence**

*Household member treated violently*

Did you see or hear a parent or household member in your home being yelled at, screamed at, sworn at, insulted or humiliated? OR Did you see or hear a parent or household member in your home being slapped, kicked, punched or beaten up? OR Did you see or hear a parent or household member in your home being hit or cut with an object?

*Physical abuse*

Did a parent, guardian or other household member spank, slap, kick, punch or beat you up? OR Did a parent, guardian or other household member hit or cut you with an object?

*Emotional abuse*

Did a parent, guardian or other household member yell, scream or swear at you, insult or humiliate you? OR Did a parent, guardian or other household member threaten to, or actually abandon you or throw you out of the house?

**Community violence**

*Bullying*

Were you bullied?

*Community violence*

Did you see or hear someone being beaten up in real life? OR Did you see or hear someone being stabbed or shot in real life? OR Did you see or hear someone being threatened with a knife or gun in real life?

*Collective violence*

Were you forced to go and live in another place due to any of these events? OR Did you experience the deliberate destruction of your home due to any of these events? OR Were you beaten up by soldiers, police, militia, or gangs? OR Was a family member or friend killed or beaten up by soldiers, police, militia, or gangs?

The sexual abuse questions were removed due to potential risks to participant and expected underreporting.

| **S2 Table. Maternal ACEs and child growth, Bachpan Cohort, Pakistan (n=877)** | | | |
| --- | --- | --- | --- |
|  | **LAZ** | **WAZ** | **WLZ** |
| *Total score* | -0.03 | 0.01 | 0.05 |
|  | (-0.08 - 0.02), 0.10 | (-0.03 - 0.06), 0.09 | (-0.01 - 0.10), 0.11 |
| *Binary* | -0.10 | 0.04 | 0.15 |
|  | (-0.27 - 0.07), 0.35 | (-0.11 - 0.19), 0.29 | (-0.02 - 0.32), 0.34 |
| *ACE categorical* |  |  |  |
| None | *ref* | *ref* | *ref* |
| One | -0.09 | 0.03 | 0.13 |
|  | (-0.32 - 0.15), 0.49 | (-0.16 - 0.22), 0.38 | (-0.07 - 0.33), 0.39 |
| Two | -0.11 | 0.11 | 0.25 |
|  | (-0.34 - 0.12), 0.46 | (-0.10 - 0.31), 0.41 | (0.00 - 0.49), 0.49 |
| Three | -0.12 | -0.07 | 0.01 |
|  | (-0.33 - 0.10), 0.44 | (-0.29 - 0.16), 0.45 | (-0.28 - 0.30), 0.57 |
| Four or more | -0.13 | 0.08 | 0.23 |
|  | (-0.42 - 0.15), 0.57 | (-0.14 - 0.30), 0.44 | (-0.06 - 0.52), 0.59 |
| *ACE Domains* |  |  |  |
| Neglect | 0.17 | 0.25 | 0.23 |
|  | (-0.21 - 0.55), 0.79 | (-0.04 - 0.53), 0.59 | (0.01 - 0.45), 0.44 |
| Psychological distress | -0.13 | -0.07 | 0.02 |
|  | (-0.32 - 0.05), 0.37 | (-0.25 - 0.12), 0.37 | (-0.19 - 0.23), 0.43 |
| Home violence | -0.07 | -0.03 | 0.02 |
|  | (-0.27 - 0.13), 0.40 | (-0.20 - 0.15), 0.36 | (-0.14 - 0.18), 0.32 |
| Community violence | -0.09 | 0.10 | 0.23 |
|  | (-0.41 - 0.23), 0.64 | (-0.15 - 0.35), 0.50 | (-0.05 - 0.52), 0.57 |
| We present β estimates for the Total score and mean differences for all other operationalizations. 95% confidence limits and confidence limit differences are also shown. We used weighted generalized linear models with cluster robust standard errors. Sampling and inverse probability censoring weights were combined. All models controlled for baseline maternal age, maternal education, trial arm, assessor, and child gender.  Abbreviations: Length-for-age z-score (LAZ); Weight-for-age z-score (WAZ); Weight-for-length z-score (WLZ) | | | |

| **S3 Table. Maternal ACEs and child development, Bachpan Cohort, Pakistan (n=877)** | | | | |
| --- | --- | --- | --- | --- |
|  | **Bayley Fine** | **Bayley Receptive** | **ASQ:SE Total** | **SDQ Total** |
| *Total score* | 0.13 | 0.22 | 1.02 | 0.14 |
|  | (-0.10 - 0.37), 0.47 | (0.08 - 0.35), 0.26 | (0.26 - 1.79), 1.50 | (-0.17 - 0.45), 0.62 |
| *Binary* | 0.44 | 0.37 | 3.30 | 0.59 |
|  | (-0.03 - 0.92), 0.96 | (0.08 - 0.65), 0.57 | (0.98 - 5.62), 4.62 | (-0.27 - 1.44), 1.70 |
| *ACE categorical* |  |  |  |  |
| None | *ref* | *ref* | *ref* | *ref* |
| One | 0.44 | 0.10 | 2.85 | 0.44 |
|  | (-0.18 - 1.05), 1.26 | (-0.20 - 0.41), 0.62 | (-0.28 - 5.98), 6.16 | (-0.65 - 1.52). 2.14 |
| Two | 0.10 | 0.48 | 3.09 | 0.86 |
|  | (-0.50 - 0.70), 1.19 | (-0.10 - 1.06), 1.15 | (-0.14 - 6.31), 6.56 | (-0.30 - 2.02), 2.35 |
| Three | 1.09 | 0.48 | 4.48 | 0.68 |
|  | (0.20 - 1.98), 1.78 | (-0.15 - 1.12), 1.28 | (0.04 - 8.92), 8.81 | (-0.59 - 1.95), 2.56 |
| Four or more | 0.33 | 1.01 | 3.86 | 0.34 |
|  | (-0.97 - 1.63), 2.58 | (0.28 - 1.75), 1.41 | (0.18 - 7.54), 7.44 | (-1.46 - 2.15), 3.61 |
| *ACE Domains* |  |  |  |  |
| Neglect | 1.35 | 0.53 | 2.28 | -0.52 |
|  | (0.57 - 2.14), 1.55 | (0.08 - 0.97), 0.89 | (-1.41 - 5.97), 7.32 | (-1.84 - 0.79), 2.64 |
| Psychological distress | 0.60 | 0.39 | 0.88 | -0.55 |
|  | (-0.30 - 1.49), 1.79 | (-0.08 - 0.86), 0.93 | (-2.08 - 3.85), 5.92 | (-1.57 - 0.46), 2.01 |
| Home violence | -0.02 | 0.11 | 1.58 | 0.99 |
|  | (-0.66 - 0.63), 1.27 | (-0.29 - 0.51), 0.80 | (-0.37 - 3.53), 3.95 | (0.20 - 1.77), 1.59 |
| Community violence | -0.55 | 0.76 | 2.95 | -0.09 |
|  | (-1.59 - 0.49), 2.13 | (0.08 - 1.44), 1.41 | (-0.67 - 6.56), 7.43 | (-1.16 - 0.99), 0.64 |
| We present β estimates for the Total score and mean differences for all other operationalizations. 95% confidence limits and confidence limit differences are also shown. We used weighted generalized linear models with cluster robust standard errors. Sampling and inverse probability censoring weights were combined. All models controlled for baseline maternal age, maternal education, trial arm, assessor, and child gender.  Abbreviations: Ages and Stages Questionnaire: Socioemotional (ASQ:SE); Strengths and Difficulties Questionnaire (SDQ). | | | | |

| **S4 Table. Maternal ACEs and child growth and development without censoring weights, Bachpan Cohort, Pakistan (n=877)** | | | | | | | |
| --- | --- | --- | --- | --- | --- | --- | --- |
|  | **LAZ** | **WAZ** | **WLZ** | **Bayley Receptive** | **Bayley Fine** | **ASQ:SE Total** | **SDQ Total** |
| *Total score* | -0.03 | 0.01 | 0.05 | 0.21 | 0.12 | 1.06 | 0.14 |
|  | (-0.08 - 0.02) | (-0.03 - 0.06) | (-0.01 - 0.10) | (0.07 - 0.34) | (-0.11 - 0.36) | (0.29 - 1.82) | (-0.17 - 0.45) |
| *Binary* | -0.11 | 0.04 | 0.15 | 0.36 | 0.43 | 3.31 | 0.60 |
|  | (-0.28 - 0.07) | (-0.11 - 0.18) | (-0.02 - 0.32) | (0.07 - 0.64) | (-0.05 - 0.91) | (1.00 - 5.62) | (-0.25 - 1.45) |
| *ACE categorical* |  |  |  |  |  |  |  |
| None | *ref* | *ref* | *ref* | *ref* | *ref* | *ref* | *ref* |
| One | -0.10 | 0.02 | 0.13 | 0.09 | 0.42 | 2.86 | 0.47 |
|  | (-0.34 - 0.15) | (-0.17 - 0.21) | (-0.07 - 0.32) | (-0.22 - 0.40) | (-0.20 - 1.05) | (-0.22 - 5.94) | (-0.60 - 1.53) |
| Two | -0.09 | 0.11 | 0.25 | 0.49 | 0.09 | 3.17 | 0.89 |
|  | (-0.32 - 0.14) | (-0.09 - 0.32) | (0.00 - 0.49) | (-0.09 - 1.06) | (-0.51 - 0.68) | (-0.11 - 6.45) | (-0.29 - 2.06) |
| Three | -0.12 | -0.08 | -0.00 | 0.50 | 1.08 | 4.29 | 0.62 |
|  | (-0.34 - 0.10) | (-0.30 - 0.15) | (-0.29 - 0.28) | (-0.14 - 1.14) | (0.19 - 1.97) | (-0.12 - 8.69) | (-0.66 - 1.90) |
| Four or more | -0.15 | 0.08 | 0.24 | 0.95 | 0.29 | 4.06 | 0.38 |
|  | (-0.43 - 0.14) | (-0.14 - 0.30) | (-0.05 - 0.54) | (0.24 - 1.65) | (-1.00 - 1.58) | (0.34 - 7.78) | (-1.43 - 2.18) |
| *ACE Domains* |  |  |  |  |  |  |  |
| Neglect | 0.17 | 0.25 | 0.23 | 0.54 | 1.35 | 2.22 | -0.61 |
|  | (-0.23 - 0.56) | (-0.05 - 0.54) | (0.01 - 0.45) | (0.09 - 0.98) | (0.57 - 2.12) | (-1.44 - 5.88) | (-1.93 - 0.71) |
| Psychological distress | -0.14 | -0.07 | 0.02 | 0.37 | 0.60 | 0.80 | -0.57 |
|  | (-0.33 - 0.04) | (-0.25 - 0.12) | (-0.19 - 0.24) | (-0.09 - 0.84) | (-0.29 - 1.50) | (-2.16 - 3.76) | (-1.58 - 0.43) |
| Home violence | -0.06 | -0.02 | 0.02 | 0.11 | -0.03 | 1.60 | 0.99 |
|  | (-0.26 - 0.14) | (-0.20 - 0.16) | (-0.14 - 0.18) | (-0.29 - 0.51) | (-0.67 - 0.60) | (-0.37 - 3.58) | (0.20 - 1.79) |
| Community violence | -0.10 | 0.10 | 0.24 | 0.72 | -0.57 | 3.11 | -0.00 |
|  | (-0.42 - 0.22) | (-0.15 - 0.35) | (-0.05 - 0.52) | (0.02 - 1.43) | (-1.63 - 0.50) | (-0.60 - 6.83) | (-1.12 - 1.11) |
| We used weighted generalized linear models with cluster robust standard errors. Sampling and inverse probability censoring weights were combined. All models controlled for baseline maternal age, maternal education, trial arm, assessor code, and child sex.  Abbreviations: Length-for-age z-score (LAZ); Weight-for-age z-score (WAZ); Weight-for-length z-score (WLZ); Ages and Stages Questionnaire: Socioemotional (ASQ:SE); Strengths and Difficulties Questionnaire (SDQ). | | | | | | | |
